# Cervical Cancer Screening Using Residual Learning

**DOI:** 10.1101/2022.12.19.22283665

**Authors:** Mohamed Abdulsamad, Esam Alsadiq Alshareef, Fawzi Ebrahim

## Abstract

Cervical cancer is a leading cancer in the female population. This disease is considered dangerous as its slow and unpredicted growth. The prevention of such cancer can be mostly achieved by screening its transformation zones. The cervical pre-cancerous zones can be considered as three types: type 1, type 2, and type 3. Screening and analyzing these three stages can be crucial for preventing their transformation into cancer. Hence, it is essentially important to have an automated and intelligent system that can grade the cervical pre-cancerous colposcopy images into one of the three types. This can help in providing the right treatment and prevent cancer transformation. In this paper, we develop a residual learning-based model (ResNet-50) to be trained for classifying the type of a colposcopy cervical image into type 1, type 2, and type 3. Experimentally, the model was fine-tuned and evaluated on a public dataset of colposcopy cervical images and achieved promising results in cervical cancer screening of accuracy of 77% and F1-score of 79%.

## 1. Introduction

Following the breast cancer, cervical cancer is the most common deadly cancer for women in America [1]. For now, such cancer is still considered uncurable in case it reaches its later stages. Moreover, it is found by the statistics of the World Health Organization (WHO) that this type of cancer is the fourth most prevalent cancer globally, with a reporting rate of 5,70,000 new cases in 2018, accounting for 7.5% of all women cancer deaths [1]. Currently, there are about 85% of the women who face death every year due to cervical cancer in the developing care where professional medical care is not well available [2].

This toll of deaths can be reduced or stopped by granting the developing world countries the access to better technology and healthcare systems in this field. This includes the availability of regular screening tests, which can help in providing an effective treatment for the cervical precancer stage. On the other hand, cervical cancer can be hard to be detected or diagnosed in its earlier stages as it has no clear and visible signs in its early stage [3]. This makes the regular screening and checkup very significant in order to prevent such disease as this earlier identification can increase the 5-years survival rate of this cancer to 66% [4].

The current methods available for screening cervical cancer are mainly the following: HPV testing, PAP smear testing, colposcopy, and biopsy [5]. The most used screening technique for the diagnosis and treatment of cervical cancer is the PAP smear. However, this technique has several drawbacks: it needs big number of microscopic examinations to for the diagnosis of cancer and noncancer patients, in addition, it is time consuming and requires trained professionals. Nevertheless, PAP smear and HPV testing are considered very expensive treatment and they provide low sensitivity in detecting cervical cancer [4].

Thus, the development of computer aid systems that can detect the precancerous stages of cervical cancer are in need. Such systems that rely only on images can be used as tools to assist medical experts in diagnosing the early stages of cervical cancer and whether or there is a probability of malignant cancer transformations.

Deep learning has shown itself as a powerful method in analyzing medical images [6]. Starting from classification to segmentation, deep learning methods have made a sharp progress in that field, making identification and detection of diseases easier and sooner.

Residual learning [7] is a deep learning method that uses identity mapping and skip connections to improve the performance of a very deep network designed to perform complex classification tasks such as cervical precancer screening.

In this paper, we investigate the capability of a residual learning-based network, denoted ResNet50, in grading the cervical precancer images into 3 stages or types: Type 1, Type 2, and Type 3. For training and testing, we used the popular cervical screening dataset [8] that consists of sufficient number of colposcopy images of the three different types.

This paper is structured as follows:

Section one is an introduction of the work, section two is review of the use of deep learning in cervical cancer screening in addition to residual learning explanation. Section three is the materials and methods. Section 4 is the results and discussion. Finally, section 5 is the conclusion.

## 1. Review

It is noticeable from the review that most of cervical cancer screening studies were conducted using hepatological or microscopic images [9-11], while a few have been conducted to diagnose cancer using colposcopy images [3,5,12]. This may be either because of the lack of the public datasets or the complexity of classifying or screening cervical pre-cancer types using colposcopy images. In general, most of the studies propose used microscopic input images in order to classify them into cancerous or non-cancerous. Other studies such as Adweb et al., [5] worked on colposcopy cervical images to classify them as normal or pre-cancerous. This can be considered as a good screening system as it may alarm medical experts that patients with pre-cancerous conditions detected are to be cared for. This can help to prevent the fast transformation into cervical cancer in later stages. This research employed residual learning-based network, ResNet50, trained with various activation function in order to select the best function for such classification task. Experimentally, this work achieved great results in terms of accuracy of diagnosing cervical cancer using normal and pre-cancerous colposcopy images.

In another study [12], a deep learning method was proposed to classify the colposcopy images into type 1, type 2, and type 3. Similar to our work, popular cervical screening dataset is used for training and testing the network presented. The authors in this work proposed a deep network named as Colposcopy Ensemble Network (CYENET) to classify cervical cancer automatically from colposcopy images. It is noticed that such deep network was capable of achieving significant results when compared to other models such as VGG16 and VGG19. When tested, the CYENET presented in this work reached an accuracy of 92.3%. However, it is important to mention that number of testing images used in this work is very low (1,884) which can be the reason this network achieves such high accuracy.

Moreover, a classification based deep learning method was proposed by Mustafa and Dauda [13]. Their method uses three different deep convolutional neural networks (DCNNs), each with different optimizers such as stochastic gradient descent (SGD), Root Mean Square Propagation (RMSprop), and Adaptive Moment Estimation (Adam), for the classification of cervical images into healthy or cancerous. This model was then trained and tested cancerous and healthy cervical images and the best optimizer based on the one that leads to the best performance of the network.

## 2. Methods

### 3.1 Dataset

In this work, we apply transfer learning approach to retrain ResNet50 to classify cervical colposcopy images into three types: 1,2, and 3. The network is trained and tested on the MobileODT Cervical Cancer Screening dataset, which is a competition launched on Kaggle [8]. The dataset contains colposcopy images of the three different types of cervical pre-cancer images (earlier stages of cervical abnormalities that may transform into cancer). Table 1 shows the number of images in each set. As seen, our learning scheme used in this work is 70:30 i.e., 70% training and 30% of images are used for testing.

**Table 1:**
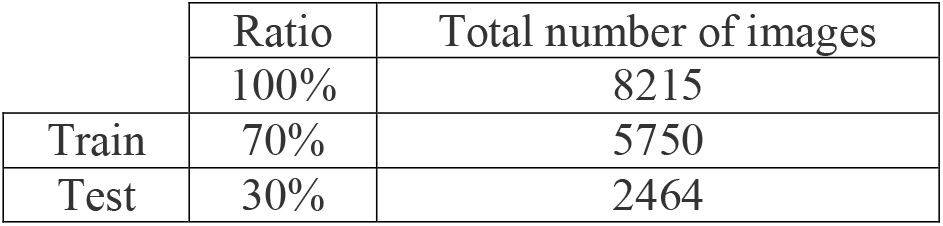
Number of train and test images

**Table 2:** Testing results

|  | Ratio | Total number of images | Testing accuracy | F1-score |
| --- | --- | --- | --- | --- |
| Test | 30% | 2464 | 77% | 79% |

Figure 1 shows a sample of the three types of cervical precancerous types.

**Figure 1:**
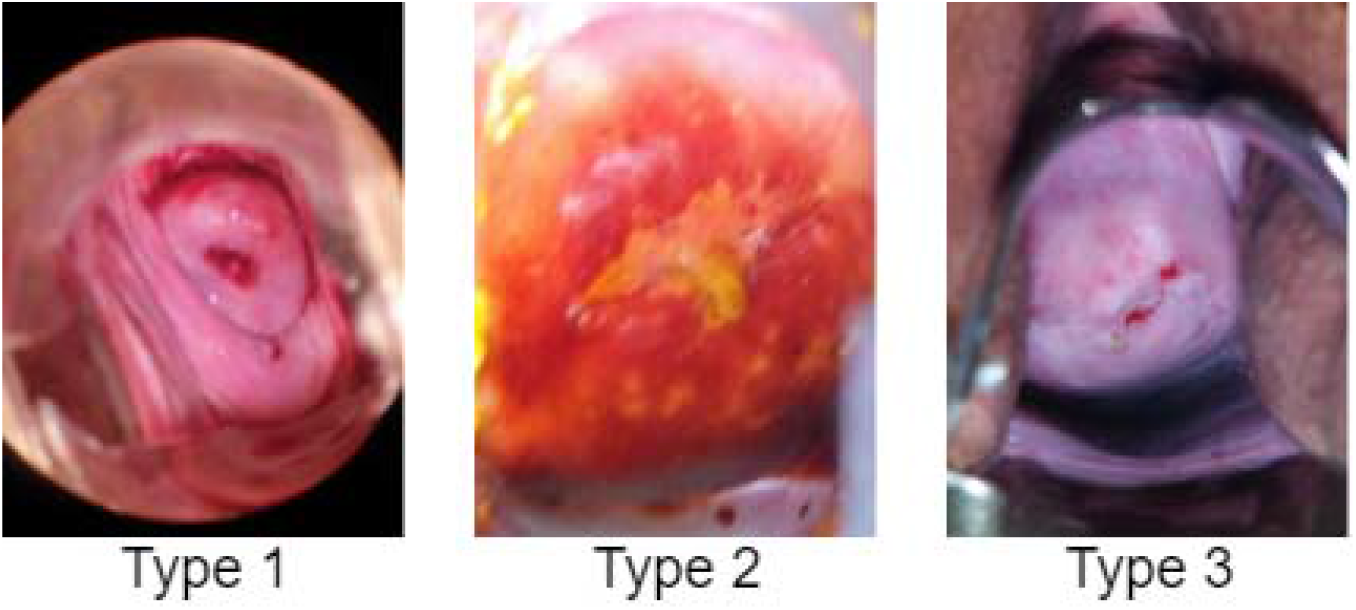
Sample of the three types of cervical colposcopy images

### 3.2 Residual Learning ResNet50

As the deep network goes deeper, it becomes more complex and sometimes it gets less effective as the increase of layers may lead to a problem called the vanishing gradient [14]. Therefore, residual learning was proposed by He et al. [15] to solve such a problem. This new learning method (ResNet) (He et al., 2016) proposed a new technique for training very deep neural networks using identity mapping for shortcut connections.

Figure 2 shows the difference between a regular deep learning block versus a residual learning block proposed in He et al. [14].

**Figure 2:**
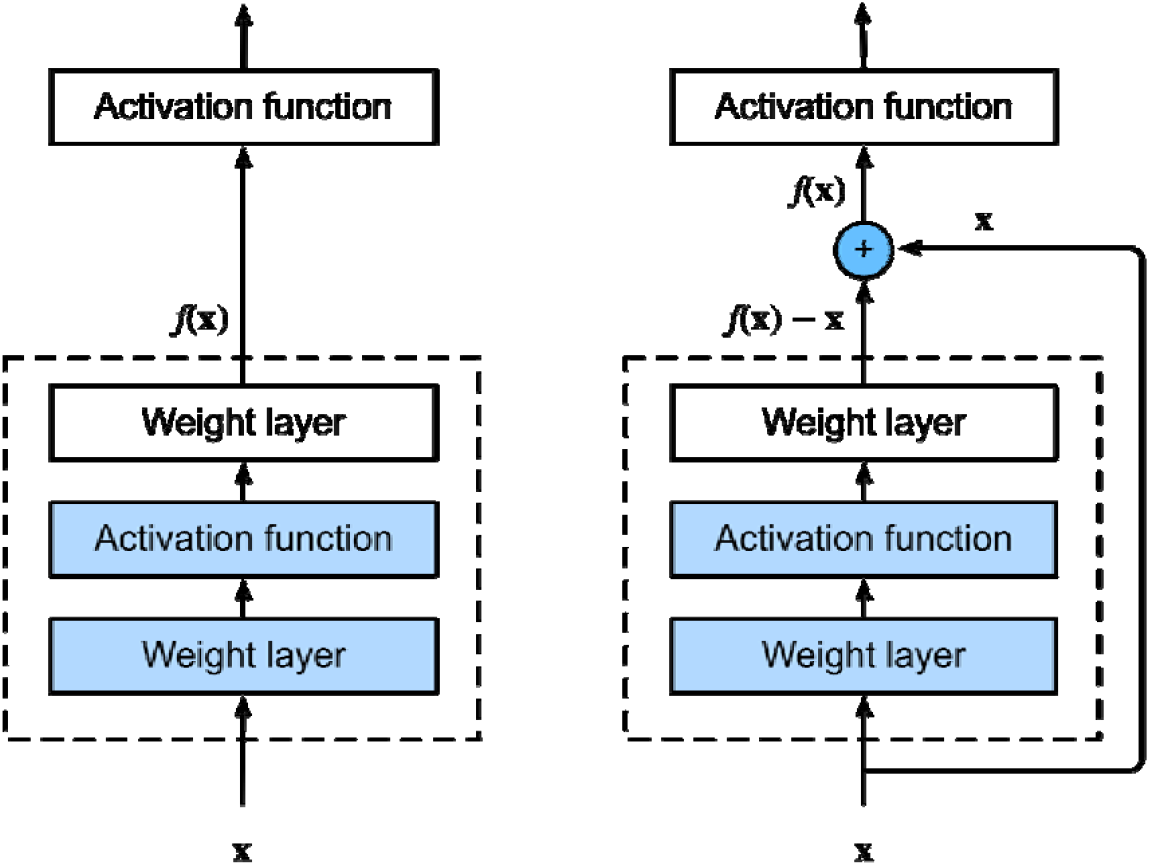
Regular (left) versus residual block (right) [15]

ResNet-50 is a residual learning-based convolutional neural network that consists of 50 layers deep. This network is originally trained on more than a million images from the ImageNet database [16]. In this paper, we adopt this pre-trained model to retrain to classify the cervical images into three types: type 1, type 2, and type 3.

To start the transfer learning process, we first used the predefined and pre-trained weights and filters for the employed ResNet-50 by providing a slight update on the learning rate in order to fine-tune the model, to fit our new cervical pre-cancer types classification.

Figure 3 illustrates the architecture of the employed transfer learning-based model to classify cervical colposcopy pre-cancerous images into three types. As seen from Figure 3, the classification part (i.e., part consisting of the fully connected layers) of the pre-trained model were removed and then replaced with three different fully connected layers that consists of two three classes: type 1, type 2, and type 3. Note that this final layer classifies cervical images based on the activations received from the previous feature extraction part which consists of convolution and pooling layers. We used 100 nodes for the first fully connected layers and 3 for last one representing the three types of cervical images. Finally, the three different activations of the last fully connected layer are fed into a Softmax layer that produces the probability of each class.

**Figure 3:**
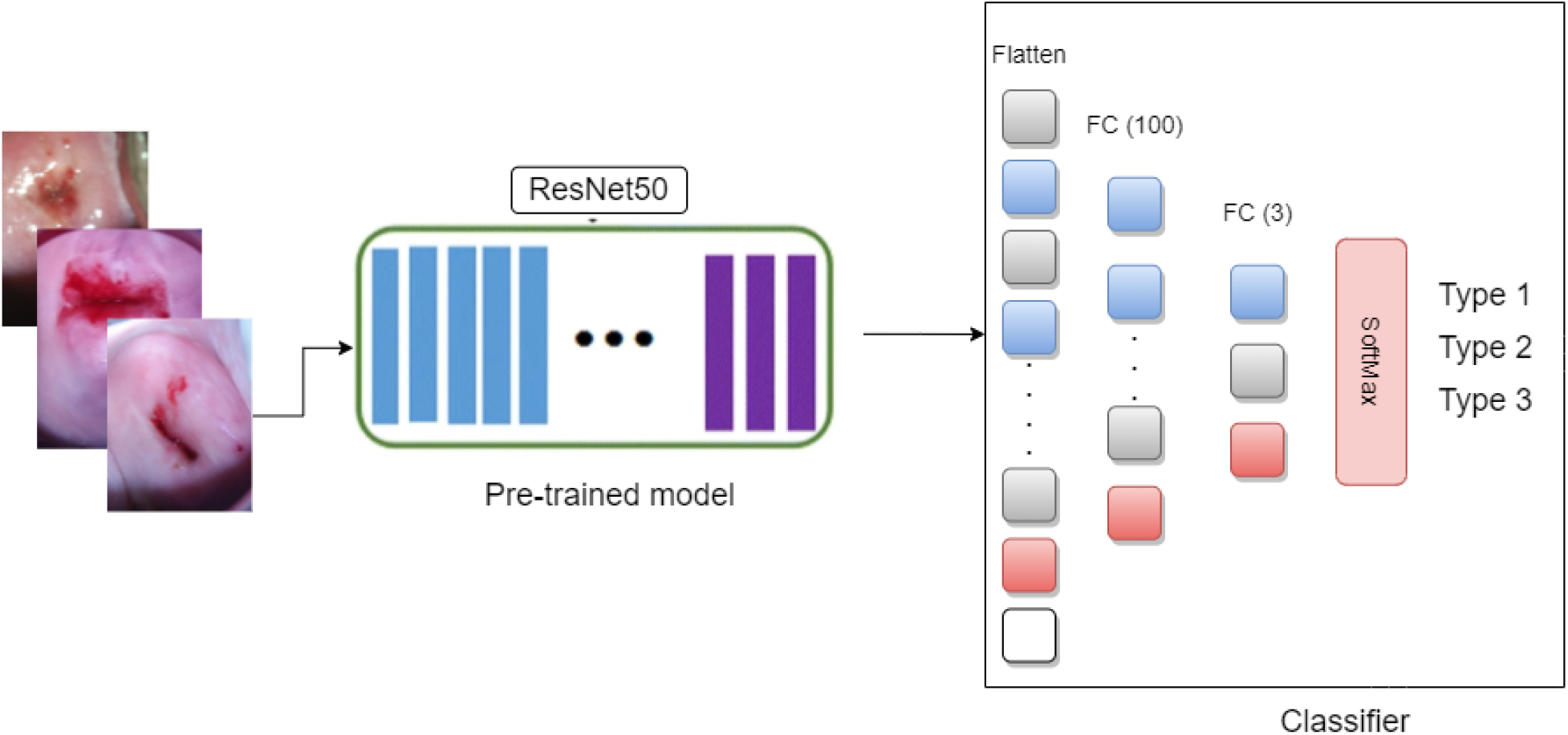
ResNet-50 schematic. Transfer learning applied to ResNet50 by replacing the last three layers with new one to fit the new cervical cancer screening classification task

## 3. Results and Discussion

### 4.1 ResNet-50 Fine-tuning

The ResNet-50 used in this work is fine-tuned using 70% of the while dataset as seen in Table 1. ResNet-50 is mainly composed of layers which includes convolution, pooling, and normalization, in addition to fully connected layers followed by a Softmax activation function in the output layer. Since we are applying transfer learning to our pre-trained model, we removed its final fully connected layer and replaces them by new layers to fit our three types of cervical cancer classification task. As for the training, we used the weights of ResNet-50 which were already learned using ImageNet dataset to extract high level features from input images. The network is retrained with a batch size of 60 images for each iteration via stochastic gradient descent [31] and a learning rate of 0.001. Figure 1 shows the training curve of the ResNet-50 when trained on 70% of the cervical colposcopy images. It is seen the error was decreasing with respect of epochs increasing until it reached a minimum value at epoch 4.

Figure 5 shows the confusion matrix of the model when tested on 30% of the images. It is seen that the model was capable of generalization accurate classification for the type 1 and 3 whereas it fails to accurately classify type 3 images. This is due to the complexity and similarity of type 2 compared to type 1 and 2 images.

**Figure 4:**
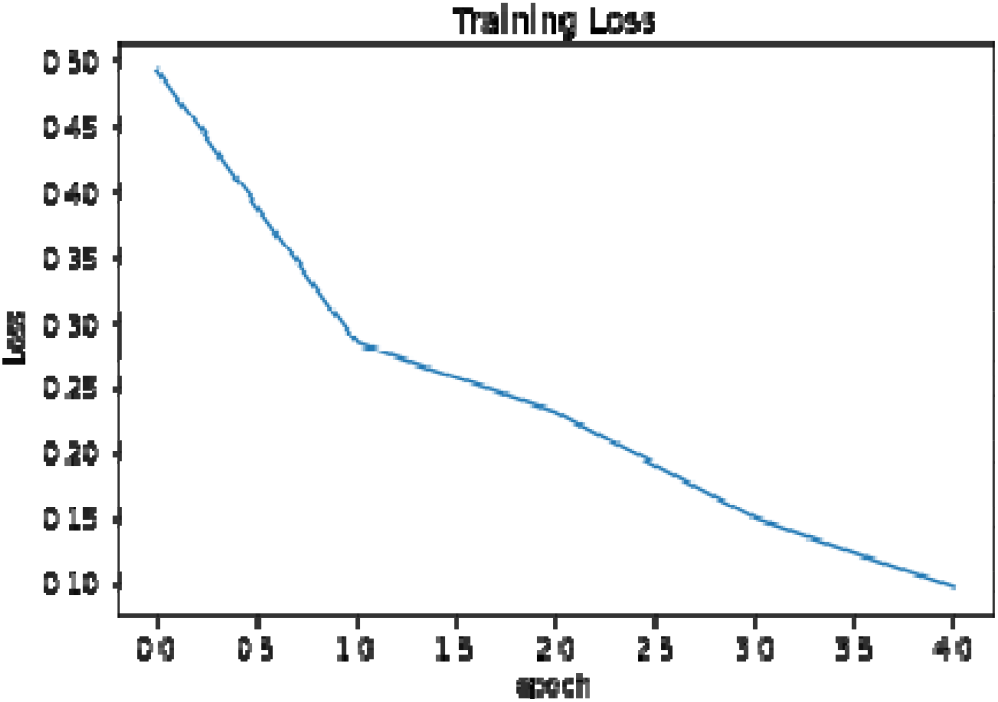
Training loss over number of epochs

**Figure 5:**
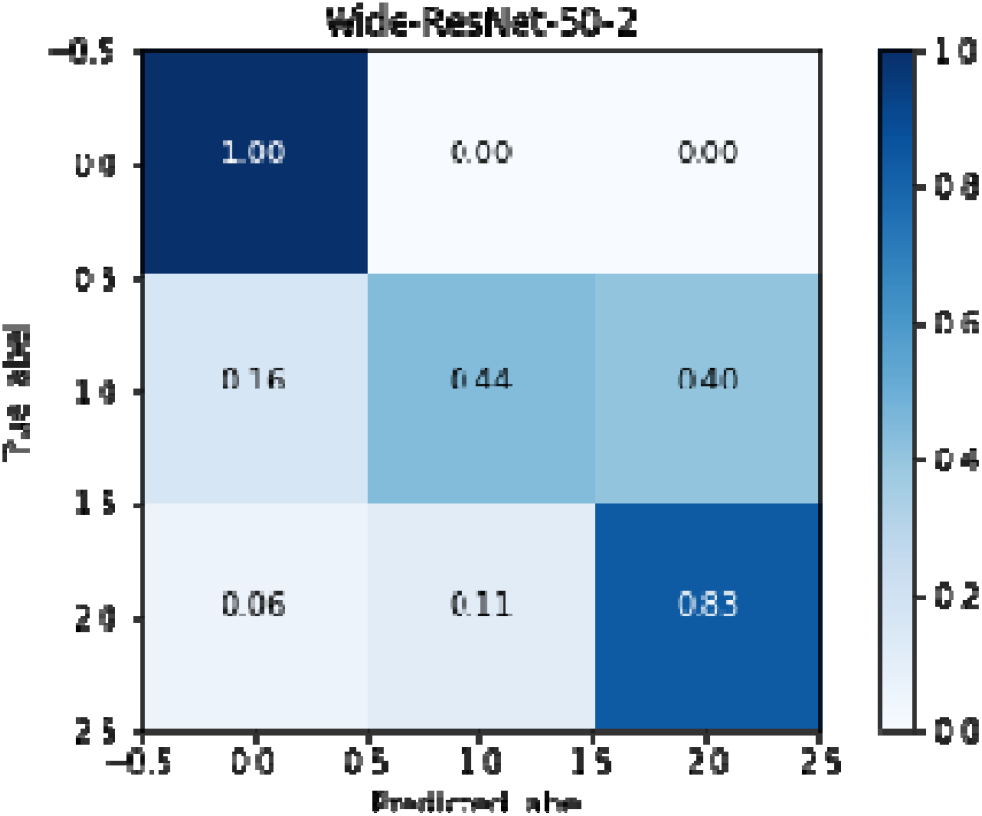
Testing confusion matrix

### 4.2 Discussion

The Resnet-50 was tested on 30% of colposcopy images which include images of the three types. As seen in figure 5 the model shows good performance in diagnosing the three different types except that it shows poorer performance in regard to type 2. One solution to increase the performance at that specific type is using data augmentation of type 2 images to increase the number of examples of that type, which may help model learn more features about type 2 class images. This can also enhance the classification accuracy of the model and helps it distinguish type 2 images from other types.

Table 4 shows the testing accuracy of the model when tested on 30% of the data. It is seen that our fine-tuned model was capable of reaching significant accuracy and F1-score of 77% and 79%, respectively.

For more understanding and analysis of the ResNet-50 model in classifying the cervical pre-cancerous stages we used Grad-Cam to visualize the areas the model relied on when predicting the type of any cervical colposcopy image. Figure 6 shows the original (first row) and activation maps (second row) of three different images of types 1, 2, and 3. As noticed, the model accurately finds some areas and distinguished features for classifying every image into one of the three different types.

**Figure 6:**
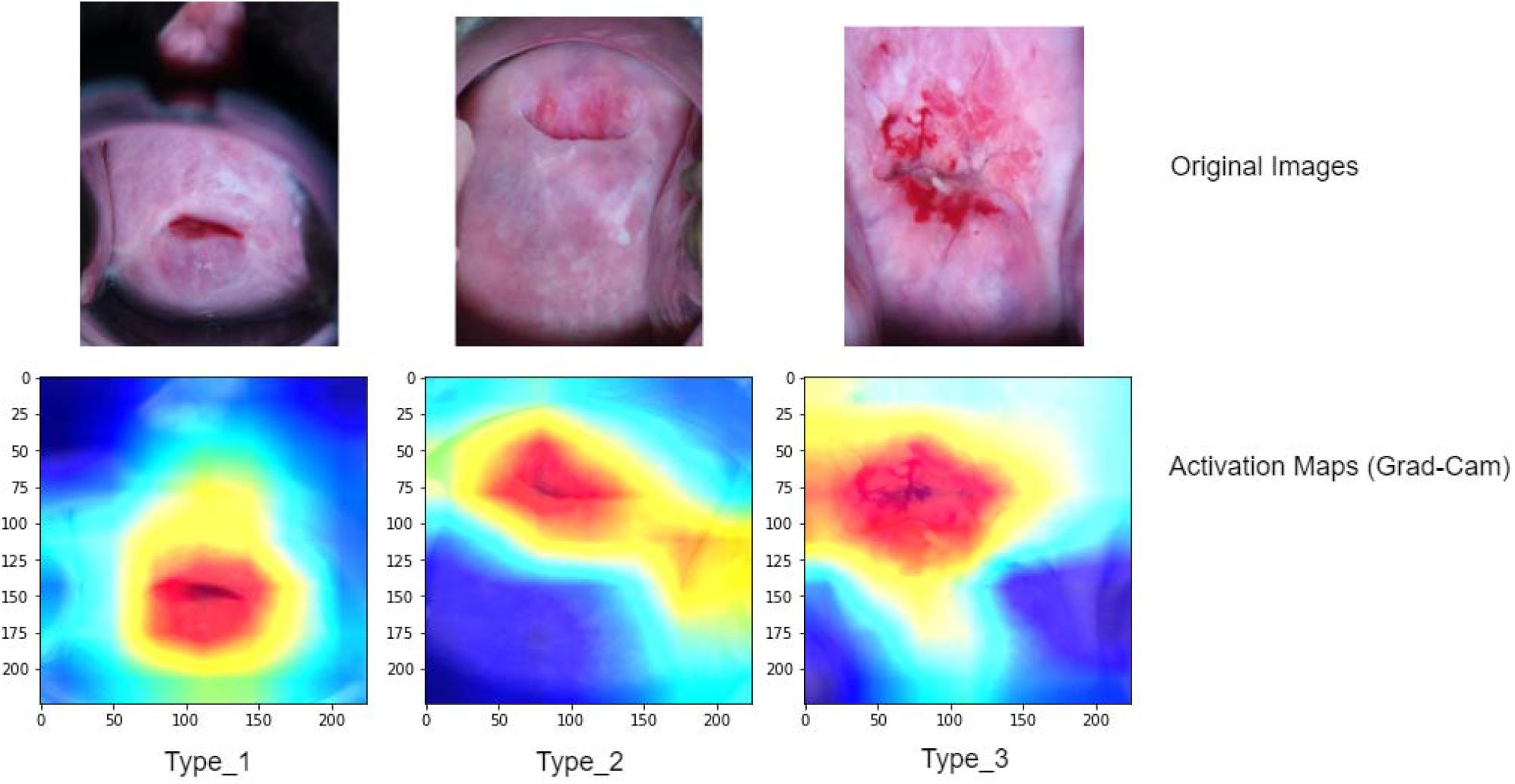
Localizations achieved by Grad-CAM technique based on the ResNet-50 model on testing cervical colposcopy images. The first row represents the original images, while the second row represents their corresponding overlaid classification activation maps.

## 4. Conclusion

In this paper, a transfer learning approach was applied to transfer the knowledge of ResNet-50 model trained on ImageNet, to perform a new classification task which is: cervical pre-cancerous colposcopy image types. The network was trained and tested on a large-scale dataset comprised of 8215 image of the three different types of cervical colposcopy images: type 1, type 2, and type 3. Experimentally, the model has shown a good performance in diagnosing the types of images, despite of the complexity of images as they are all quite similar and contain some artefacts such as blood and others.

It is concluded that ResNet-50 is a well-designed and deep architecture of sufficient complexity, could achieve significantly higher classification accuracy when distinguishing between these three types of cervical pre-cancerous colposcopy images which helps in better screening. Furthermore, it is noticed that this model learned features and activation maps of every type, as it was shown in the Grad-Cam visualization which demonstrates that mid and high-level features are learned effectively by the model.

Overall, it can be stated that such deep model can be used in real-life application and a helping tool for cervical pre-cancer stages screening in addition to their visualization.

## Data Availability

All data produced in the present study are available upon reasonable request to the authors.

